# Pre-Existing Traits Associated with Covid-19 Illness Severity

**DOI:** 10.1101/2020.04.29.20084533

**Authors:** Joseph E. Ebinger, Natalie Achamallah, Hongwei Ji, Brian L. Claggett, Nancy Sun, Patrick Botting, Trevor-Trung Nguyen, Eric Luong, Elizabeth H. Kim, Eunice Park, Yunxian Liu, Ryan Rosenberry, Yuri Matusov, Steven Zhao, Isabel Pedraza, Tanzira Zaman, Michael Thompson, Koen Raedschelders, Anders H. Berg, Jonathan D. Grein, Paul W. Noble, Sumeet S. Chugh, C. Noel Bairey Merz, Eduardo Marbán, Jennifer E. Van Eyk, Scott D. Solomon, Christine M. Albert, Peter Chen, Susan Cheng

**Affiliations:** Department of Cardiology, Cedars-Sinai Medical Center, Los Angeles, California, USA; Smidt Heart Institute, Cedars-Sinai Medical Center, Los Angeles, California, USA; Department of Medicine, Cedars-Sinai Medical Center, Los Angeles, California, USA; Division of Pulmonary and Critical Care Medicine, Cedars-Sinai Medical Center, Los Angeles, California, USA; Shanghai Tenth People’s Hospital, Tongji University, Shanghai, China; Cardiovascular Division, Brigham and Women’s Hospital, Boston, Massachusetts, USA; Enterprise Information Systems Data Intelligence Team, Cedars-Sinai Medical Center, Los Angeles, California, USA; Advanced Clinical Biosystems Institute, Cedars-Sinai Medical Center, Los Angeles, California, USA; Department of Pathology and Laboratory Medicine, Cedars-Sinai Medical Center, Los Angeles, California, USA; Department of Epidemiology, Cedars-Sinai Medical Center, Los Angeles, California, USA; Women’s Guild Lung Institute, Cedars-Sinai Medical Center, Los Angeles, California, USA; Barbra Streisand Women’s Heart Center, Cedars-Sinai Medical Center, Los Angeles, California, USA

## Abstract

**Background:** Certain individuals, when infected by SARS-CoV-2, tend to develop the more severe forms of Covid-19 illness for reasons that remain unclear.

**Methods:** We studied N=442 patients who presented with laboratory confirmed Covid-19 illness to our U.S. metropolitan healthcare system. We curated data from the electronic health record, and used multivariable logistic regression to examine the association of pre-existing traits with a Covid-19 illness severity defined by level of required care: need for hospital admission, need for intensive care, and need for intubation.

**Results:** Of all patients studied, 48% required hospitalization, 17% required intensive care, and 12% required intubation. In multivariable-adjusted analyses, patients requiring a higher levels of care were more likely to be older (OR 1.5 per 10 years, P<0.001), male (OR 2.0, P=0.001), African American (OR 2.1, P=0.011), obese (OR 2.0, P=0.021), with diabetes mellitus (OR 1.8, P=0.037), and with a higher comorbidity index (OR 1.8 per SD, P<0.001). Several clinical associations were more pronounced in younger compared to older patients (P_interaction_<0.05). Of all hospitalized patients, males required higher levels of care (OR 2.5, P=0.003) irrespective of age, race, or morbidity profile.

**Conclusions:** In our healthcare system, greater Covid-19 illness severity is seen in patients who are older, male, African American, obese, with diabetes, and with greater overall comorbidity burden. Certain comorbidities paradoxically augment risk to a greater extent in younger patients. In hospitalized patients, male sex is the main determinant of needing more intensive care. Further investigation is needed to understand the mechanisms underlying these findings.

## INTRODUCTION

The severe acute respiratory syndrome coronavirus-2 (SARS-CoV-2) is now well recognized as the cause of the coronavirus disease 2019 (Covid-19) global pandemic.^1–3^ The rate of rise in Covid-19 infection and its associated outcomes in the United States is now comparable to rates observed in other severely affected countries such as China, Italy, and Spain.^4–10^ The spread of Covid-19 in the United States has been especially pronounced in the states of California, New York, Michigan, Louisiana, and Washington.^11^ Consistently reported across all regions is the observation that, of all individuals who become infected with SARS-CoV-2, a majority tend to have mild or no symptoms; however, an important minority will develop predominantly respiratory disease that can lead to critical illness and death.^12–15^ Multiple, reports suggest that certain demographic and clinical characteristics may predispose infected persons to more severe manifestations of Covid-19, such as older age, male sex, and pre-existing hypertension, pulmonary disease, or cardiovascular disease.^4,16–20^ Given that these traits tend to cluster among the same persons, the relative contribution of each trait to the risk for developing more severe presentations of Covid-19 illness remains unclear.

We conducted a comprehensive investigation of the pre-existing demographic and clinical correlates of Covid-19 illness severity observed among patients evaluated for Covid-19 within our multi-site healthcare system in Los Angeles, California. We deliberately focused our study on pre-existing characteristics for two main reasons: first, we recognize that patients with Covid-19 illness can present early or late in the disease course, causing many clinical features to vary at the time of initial clinical encounter; and, second, we anticipate that ongoing public health efforts can be informed and augmented by understanding which predisposing factors may render certain segments of the population at higher risk for the most morbid sequelae of SARS-CoV-2 infection.

## METHODS

### Study Sample

The Cedars-Sinai Health System is located in Los Angeles, California with a diverse catchment area of 1.8 million individuals, 33% of whom are over the age of 45 years and 80% identify as a racial or ethnic minority. The Cedars-Sinai Health System includes Cedars-Sinai Medical Center (CSMC), Marina Del Rey Hospital (MDRH), and affiliated clinics. For the current study, we included all patients who were found to have a laboratory confirmed diagnosis of SARS-CoV-2 infection while being evaluated or treated for signs or symptoms concerning for Covid-19 at CSMC or MDRH, beginning after the first confirmed case of community transmission was reported in the U.S. on Feburary 26, 2020. Subsquently, the first laboratory confirmed Covid-19 case in our health system was on March 8, 2020. All laboratory testing for SARS-CoV-2 has been performed using reverse transcriptase polymerase chain reaction of extracted RNA from nasopharyngeal swabs. All patient testing was performed by the Los Angeles Department of Public Health until March 21, 2020, at which time the CSMC Department of Pathology and Laboratory Medicine began using the A*STAR FORTITUDE KIT 2.0 COVID-19 Real-Time RT-PCR Test (Accelerate Technologies Pte Ltd, Singapore). For the minority of patients in our study who had SARS-CoV-2 testing performed at an outside facility (3.6%), documentation of a positive test was carefully reviewed by our medical staff and considered comparable for accuracy.

### Data Collection

For all patients considered to have Covid-19, based on direct or documented laboratory test result and suggestive signs and/or symptoms, we obtained data from the electronic health record (EHR) on the following demographic and clinical characteristics: age at the time of diagnosis, sex, race, ethnicity, body mass index (BMI), smoking status, comorbidities (as coded by ICD-10), and vital signs and laboratory diagnostics assessed within 1 week prior to presentation. We conducted iterative quality control and quality assurance analyses on all data extracted directly from the EHR and used manual chart review to verify collected data on clinical characteristics where appropriate. To capture variation in relative comorbid status, in a way that is not captured by distinct medical history variables alone, we calculated the Elixhauser Comorbidity Index (ECI) with van Walraven weighting for all patients based on all available clinical data.^21–24^ The ECI uses 31 categories to quantify a patient’s burden of comorbid conditions and has been shown to outperform other indices in predicting adverse outcomes (**Supplemental Table 1**).^23–29^ For patients admitted to the hospital, length of stay, admission to an intensive care unit (ICU) and death were ascertained from time stamps recorded for admission, unit transfers, and discharge. Interventions such as intubation and prone positioning were identified through time stamped orders in the EHR and verified by manual chart review. Dates and times of onset for reported or observed relevant signs and/or symptoms were also determined via manual chart review. All care was provided at the discretion of the treating physicians. Our outcomes for this study included: *severe illness* (defined as requiring any kind of hospital admission), *critical illness* (defined as the need for intensive care during hospitalization), and *respiratory failure* (defined as the need for intubation and mechanical ventilation). The CSMC institutional review board approved all protocols for the current study.

### Statistical Analyses

For the total sample of Covid-19 patients, we used parametric tests to compare normally distributed continuous variables and non-normally distributed or categorical variables, respectively. We also used histograms to display age and sex distribution for the total cohort, the patients admitted but not requiring intensive care, and patients requiring intensive care at any time during hospitalization, and the patient requiring intubation and mechanical ventilation at any time during hospitalization. We used ordinal logistic regression to examine the associations between pre-existing characteristics (based on clinically relevant, non-missing data) and a primary outcome measure of illness severity, defined as an illness severity score. We constructed the illness severity score, with higher values assigned to needing more intensive levels of clinical care, based on the following stepwise categories: 0 = clinically deemed to not require admission; 1 = required hospital admission but never required intensive care; 2 = required intensive level care but never intubation; and, 3 = required intubation during hospitalization. We constructed age- and sex-adjusted models, from which significantly associated covariates (based on P<0.20) were selected for inclusion in the final multivariable-adjusted models, where appropriate (i.e. smaller sample sizes). In secondary analyses, we analyzed the associations of pre-existing patient characteristics with the distinct outcomes of needing any hospital admission (severe illness) and, in the cohort of all hospitalized patients representing an especially vulnerable population, the need for intensive care (critical illness) or intubation (respiratory failure). All analyses were performed using R, version 3.5.1 (R Foundation for Statistical Computing) and Stata, version 15 (StataCorp). For all final models, P values were 2-sided and considered significant at threshold level of 0.05.

## RESULTS

Regional analyses showed that patients presented to our healthcare system from across a broad geographic catchment area in Los Angeles County (**Supplemental Figure 1**). The demographic and clinical characteristics of all patients in our study sample are shown in **Table 1**. Overall, almost half of patients (N=214, 48%) were clinically assessed to require hospital admission, of whom over a third (N=77; 36%) required intensive care and almost a quarter (24.3%) required intubation. In unadjusted analyses, the patients who were more likely to require higher levels of care tended to be older, male, African American, and with known obesity, hypertension, diabetes mellitus, higher Elixhauser comorbidity index, and have prior myocardial infarction or heart failure (**Table 1**). The number of men with confirmed Covid-19 infection outnumbered women in nearly all age groups; this sex difference was more pronounced among patients requiring hospitalization and particularly among patients requiring intensive care or intubation (**Figure 1**). We also observed a consistently higher rate of greater illness severity among African Americans compared to persons of other racial groups (**Figure 1**).

**Table 1.** Demographic and Clinical Characteristics of All Patients With Covid-19.

|  | Total | Covid-19 Illness Severity |  |  | *P* value* |
| --- | --- | --- | --- | --- | --- |
|  |  | *Not*<br>*Admitted* | *Admitted,*<br>*Non-ICU* | *Admitted,*<br>*ICU* |  |
| N | 442 | 228 | 137 | 77 |  |
| Age, years, mean ± sd | 52.7 ± 19.7 | 43.2 ± 15.5 | 61.7 ± 19.5 | 65.1 ± 16.3 | <0.001 |
| Male sex, n (%) | 256 (58) | 121 (53) | 78 (57) | 57 (74) | 0.005 |
| Ethnicity, n (%) |  |  |  |  |  |
| Non-Hispanic | 341 (77) | 167 (73) | 112 (82) | 62 (81) | 0.003 |
| Hispanic | 68 (15) | 33 (15) | 22 (16) | 13 (17) |  |
| Unknown | 33 (8) | 28 (12) | 3 (2) | 2 (3) |  |
| Race, n (%) |  |  |  |  |  |
| White | 283 (64) | 136 (60) | 99 (72) | 48 (62) | <0.001 |
| African American | 58 (13) | 19 (8) | 21 (15) | 18 (23) |  |
| Asian | 35 (8) | 25 (11) | 7 (5) | 3 (4) |  |
| Other | 37 (8) | 23 (10) | 8 (6) | 6 (8) |  |
| Unknown | 29 (7) | 25 (11) | 2 (2) | 2 (3) |  |
| Obesity, n (%) | 71 (16) | 27 (12) | 27 (20) | 17 (22) | 0.04 |
| Hypertension, n (%) | 161 (36) | 44 (19) | 68 (50) | 49 (64) | <0.001 |
| Diabetes mellitus, n (%) | 84 (19) | 18 (8) | 40 (29) | 26 (34) | <0.001 |
| Elixhauser comorbidity score, mean ± sd | 6.32 ± 10.8 | 1.31 ± 4.4 | 10.62 ± 12.2 | 13.5 ± 13.8 | <0.001 |
| Prior myocardial infarction or heart failure, n (%) | 49 (11) | 4 (2) | 27 (20) | 18 (23) | <0.001 |
| Prior COPD or asthma, n (%) | 70 (16) | 27 (12) | 28 (20) | 15 (20) | 0.059 |
| ACE inhibitor use, n (%) | 31 (7) | 11 (5) | 15 (11) | 5 (7) | 0.084 |
| Angiotensin receptor blocker use, n (%) | 41 (9) | 13 (6) | 15 (11) | 13 (17) | 0.01 |

**Figure 1.**
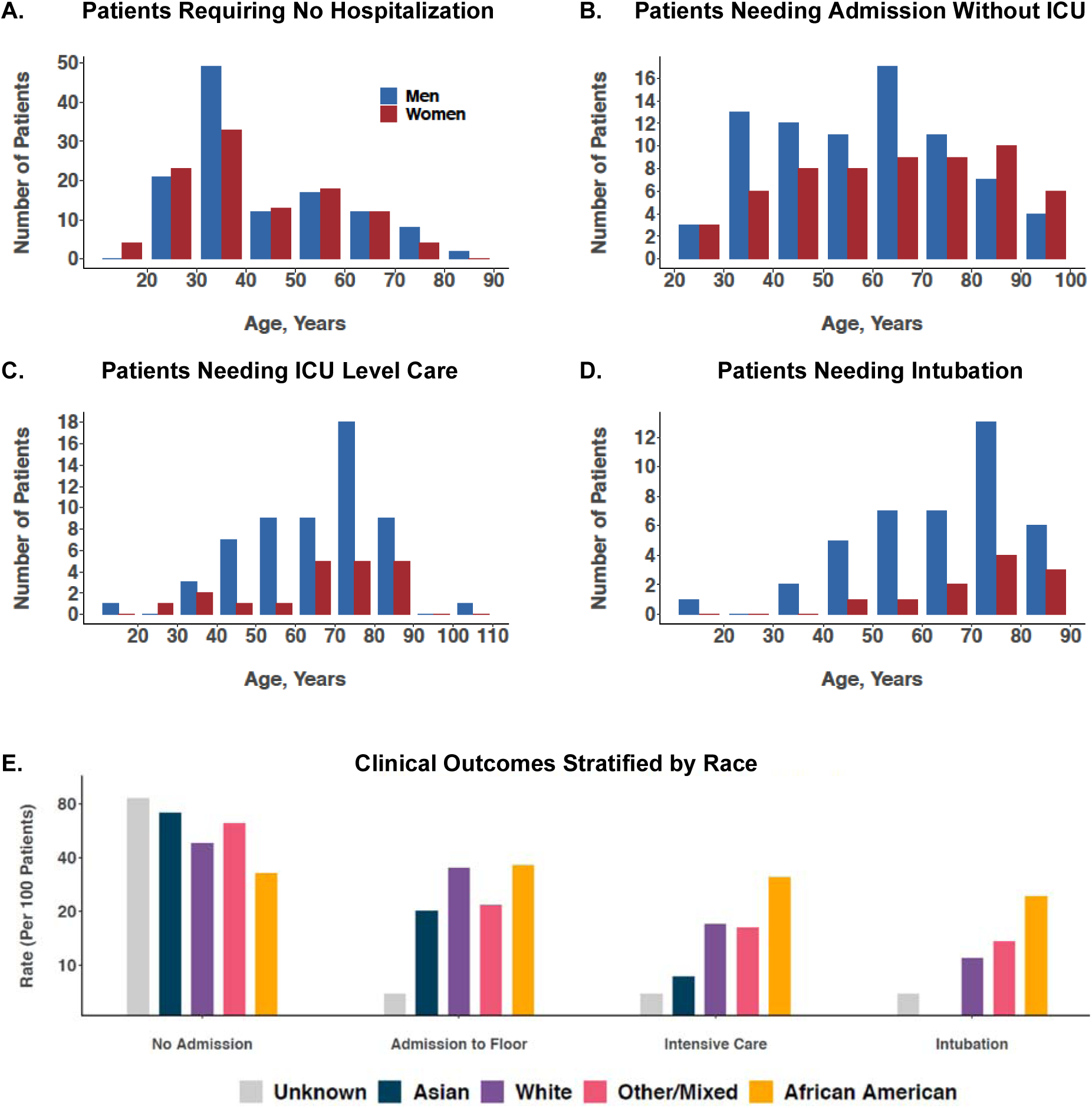
Age, Sex, and Race Distributions of Patients with Covid-19, Stratified by Admission Status. The frequency of laboratory confirmed Covid-19 was higher in males compared to females particularly among individuals requiring hospital admission, individuals with critical illness (requiring intensive care), and individuals with respiratory failure (requiring intubation). The frequency of African Americans manifesting more severe forms of Covid-19 illness, requiring higher levels of clinical care, was greater than that for other racial groups.

For the primary outcome of illness severity, categorized by escalating levels of care (i.e., hospitalization, intensive care, intubation), the pre-existing characteristics that demonstrated statistical significance in age- and sex-adjusted models included older age, male sex, African American race, obesity, hypertension, diabetes mellitus, and the Elixhauser comorbidity score (**Table 2, Figure 2**). The associations that remained significant in the fully-adjusted multivariable model included older age (odds ratio [OR] 1.49 per 10 years, 95% confidence interval [CI] 1.30-1.70, P<0.001), male sex (OR 2.01, 95% CI 1.34–3.04, P=0.001), African American race (OR 2.13, 95% CI 1.19–3.83, P=0.011), obesity (OR 1.95, 95% CI 1.11–3.42, P=0.021), diabetes mellitus (OR 1.77, 95% CI 1.03–3.03, P=0.037) and the comorbidity score (OR 1.77 per SD, 95% CI 1.37–2.28, P<0.001). We also observed a trend towards lower severity of illness among patients chronically treated with angiotensin converting enzyme (ACE) inhibitor therapy, with OR 0.48 (95% CI 0.22–1.04; P=0.06).

**Table 2.**
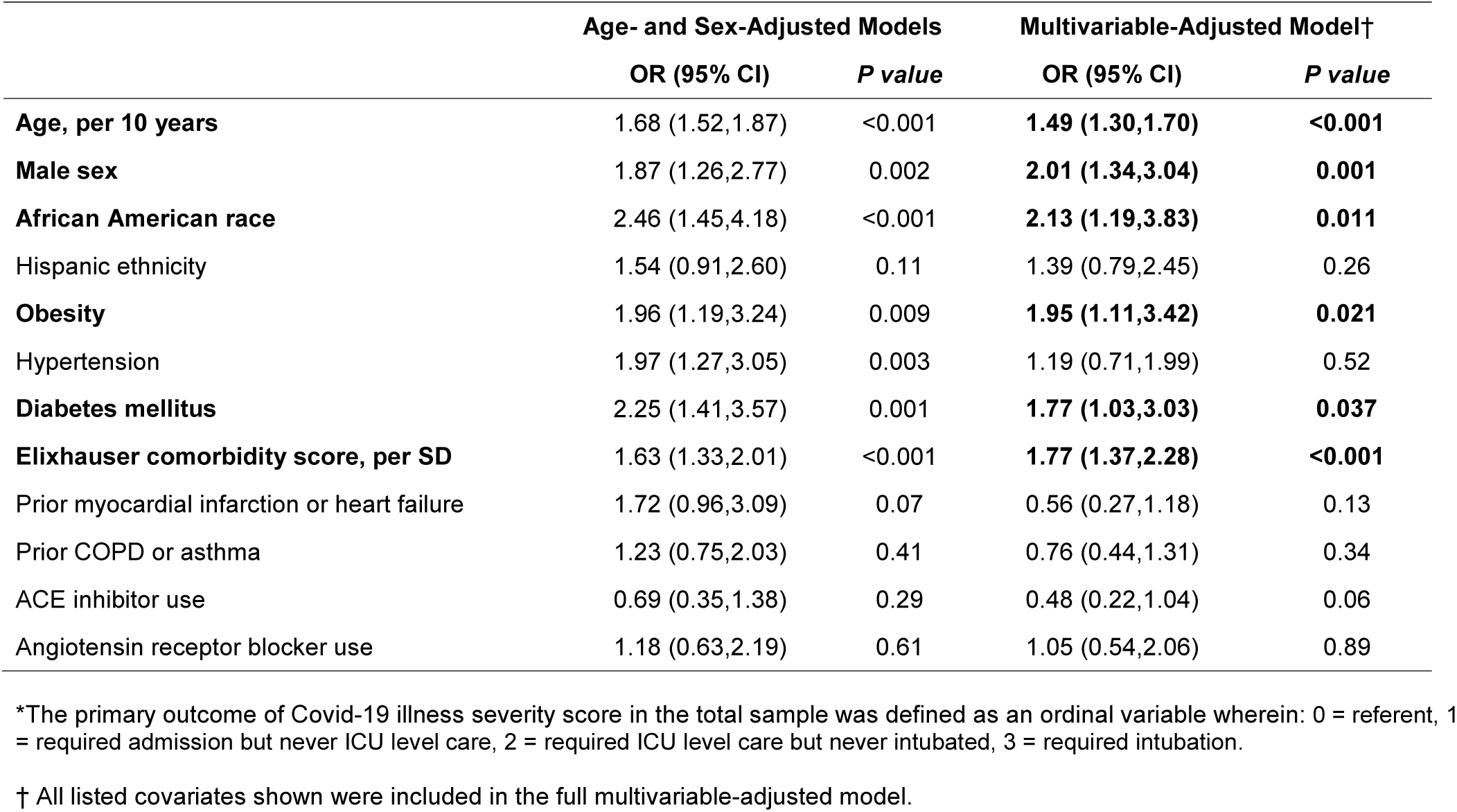
Characteristics Associated with Overall Covid-19 Illness Severity* in the Total Sample (N=442)

|  | Age- and Sex-Adjusted Models |  | Multivariable-Adjusted Model† |  |
| --- | --- | --- | --- | --- |
|  | OR (95% CI) | *P* value | OR (95% CI) | *P* value |
| **Age, per 10 years** | 1.68 (1.52,1.87) | <0.001 | **1.49 (1.30,1.70)** | **<0.001** |
| **Male sex** | 1.87 (1.26,2.77) | 0.002 | **2.01 (1.34,3.04)** | **0.001** |
| **African American race** | 2.46 (1.45,4.18) | <0.001 | **2.13 (1.19,3.83)** | **0.011** |
| Hispanic ethnicity | 1.54 (0.91,2.60) | 0.11 | 1.39 (0.79,2.45) | 0.26 |
| **Obesity** | 1.96 (1.19,3.24) | 0.009 | **1.95 (1.11,3.42)** | **0.021** |
| Hypertension | 1.97 (1.27,3.05) | 0.003 | 1.19 (0.71,1.99) | 0.52 |
| **Diabetes mellitus** | 2.25 (1.41,3.57) | 0.001 | **1.77 (1.03,3.03)** | **0.037** |
| **Elixhauser comorbidity score, per SD** | 1.63 (1.33,2.01) | <0.001 | **1.77 (1.37,2.28)** | **<0.001** |
| Prior myocardial infarction or heart failure | 1.72 (0.96,3.09) | 0.07 | 0.56 (0.27,1.18) | 0.13 |
| Prior COPD or asthma | 1.23 (0.75,2.03) | 0.41 | 0.76 (0.44,1.31) | 0.34 |
| ACE inhibitor use | 0.69 (0.35,1.38) | 0.29 | 0.48 (0.22,1.04) | 0.06 |
| Angiotensin receptor blocker use | 1.18 (0.63,2.19) | 0.61 | 1.05 (0.54,2.06) | 0.89 |

\*The primary outcome of Covid-19 illness severity score in the total sample was defined as an ordinal variable wherein: 0 = referent, 1 = required admission but never ICU level care, 2 = required ICU level care but never intubated, 3 = required intubation.
† All listed covariates shown were included in the full multivariable-adjusted model.

**Figure 2.**
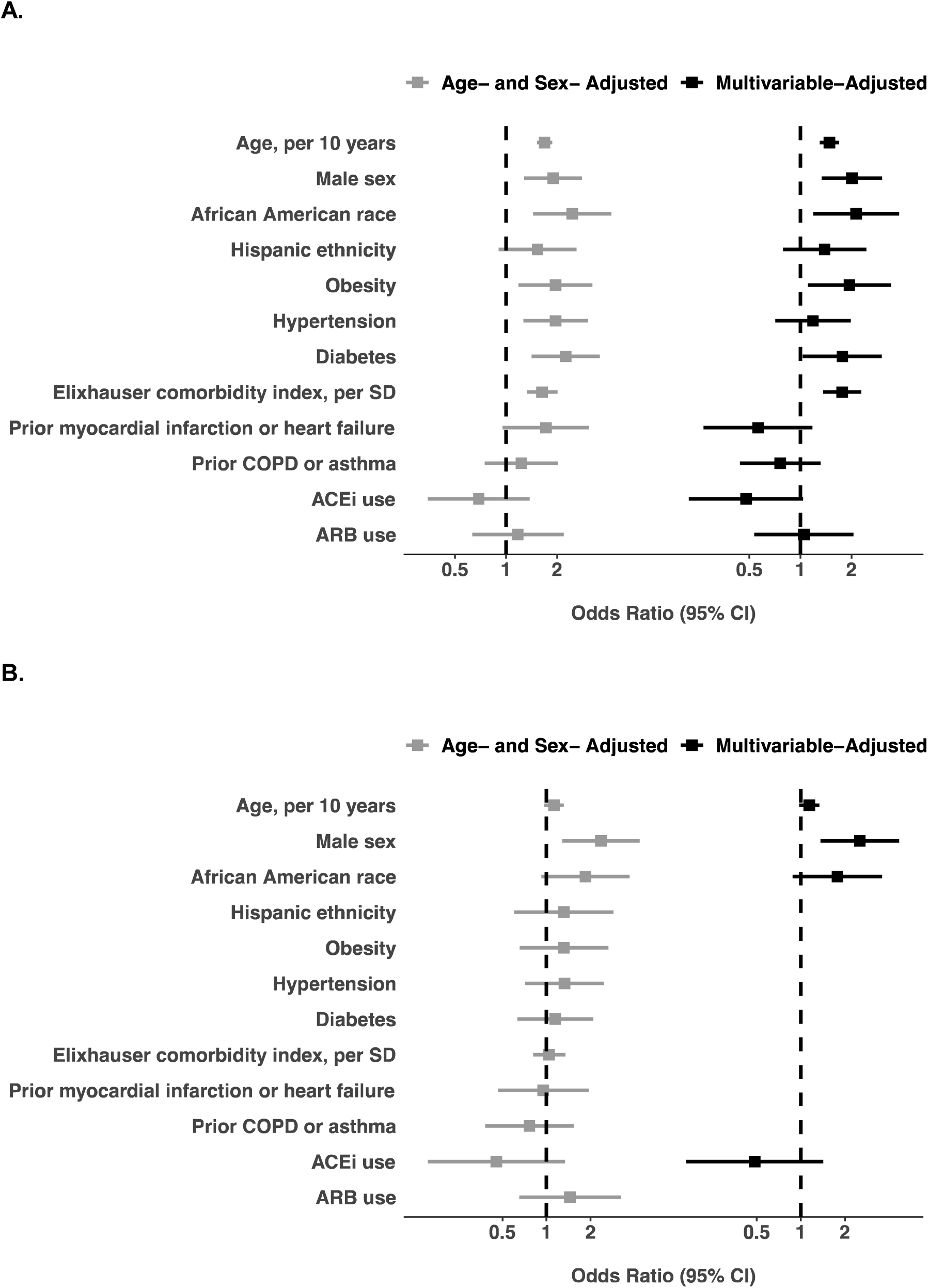
Characteristics Associated with Overall Covid-19 Illness Severity. The primary outcome of Covid-19 illness severity score was defined as an ordinal variable wherein: 0 = referent, 1 = required admission but never ICU level care, 2 = required ICU level care but never intubate, 3 = required intubation. Results for the total sample of N=442 admitted and nonadmitted patients are shown in **Panel A** (all listed covariates shown were in the full multivariable-adjusted model). Results for the N=214 admitted patients are shown in **Panel B** (to avoid model overfitting given the smaller sample size, covariates included in the multivariable model were selected from age- and sex-adjusted models based on significance with P<0.20).

For the specific outcome of needing any hospital admission, the pre-admission characteristics that demonstrated statistical significance included older age, male sex, African American race, obesity, hypertension, diabetes mellitus, the Elixhauser comorbidity index, and prior myocardial infarction or heart failure (**Supplemental Table 2**). In the multivariable model adjusting for all key covariates, the pre-existing traits that remained significantly associated with needing any hospital admission were older age, diabetes mellitus, and higher comorbidity index.

Among the patients whose illness severity required hospitalization, male sex was associated with the outcome of requiring further escalating levels of care (i.e., intensive care and intubation) (**Table 3**). In the multivariable model adjusting for key covariates, male sex remained the single most important risk marker of requiring higher-level care (OR 2.53, 95% CI 1.36–4.70, P=0.003). The results for male sex were similar for the individual outcomes of requiring intensive care or intubation (**Supplemental Table 3**). We again observed a trend towards lower need for intensive care among patients chronically taking an ACE inhibitor (OR 0.38, 95% CI 0.13–0.17, P=0.09), and greater need for intubation among African Americans patients (OR 2.14, 95% CI 0.99–4.64, P=0.053).

**Table 3.** Characteristics Associated with Covid-19 Illness Severity Among All Hospitalized Patients.

|  | Age- and Sex-Adjusted Models |  | Multivariable-Adjusted Model† |  |
| --- | --- | --- | --- | --- |
|  | OR (95% CI) | *P* value | OR (95% CI) | *P* value |
| Age, per 10 years | 1.13 (0.97,1.32) | 0.12 | 1.14 (0.98,1.34) | 0.09 |
| **Male sex** | 2.36 (1.29,4.34) | 0.006 | **2.53 (1.36,4.70)** | **0.003** |
| African American race | 1.85 (0.92,3.71) | 0.08 | 1.78 (0.88,3.58) | 0.11 |
| Hispanic ethnicity | 1.31 (0.60,2.86) | 0.49 | - | - |
| Obesity | 1.32 (0.66,2.65) | 0.44 | - | - |
| Hypertension | 1.33 (0.72,2.46) | 0.37 | - | - |
| Diabetes mellitus | 1.15 (0.63,2.09) | 0.65 | - | - |
| Elixhauser comorbidity score, per SD | 1.04 (0.81,1.34) | 0.74 | - | - |
| Prior myocardial infarction or heart failure | 0.95 (0.47,1.94) | 0.89 | - | - |
| Prior COPD or asthma | 0.77 (0.38,1.54) | 0.46 | - | - |
| ACE inhibitor use | 0.45 (0.15,1.34) | 0.15 | 0.48 (0.16,1.43) | 0.19 |
| Angiotensin receptor blocker use | 1.45 (0.65,3.21) | 0.36 | - | - |

\*The secondary outcome of Covid-19 illness severity score in hospitalized patients was defined as an ordinal variable wherein: 1 = referents required admission but never ICU level care, 2 = required ICU level care but never intubate, 3 = required intubation.
†To avoid model overfitting given the sample size, covariates included in the multivariable model were selected from age- and sex-adjusted models based on significance with $P < 0.20$ .

In secondary analyses, we used multiplicative interaction terms to assess for effect modification for associations observed in the main analyses (**Supplemental Table 4**). While considered exploratory or hypothesis generating analyses, we found several interactions of potential interest (**Figure 3**). In particular, the associations of Hispanic ethnicity, obesity, diabetes, and Elixhauser comorbidity index with the primary outcome appeared paradoxically more pronounced in younger compared to older individuals (**Supplemental Table 5**). By contrast, the primary outcome was more prounounced among older compared to younger African Americans. Also paradoxically, hypertension appeared associated with greater risk in non-obese patient and with lower risk in obese patients. We repeated all main analyses with additional adjustment for smoking status in the subset of patients with available data on smoking; in these models, all signicant results remained unchanged (data not shown).

**Figure 3.**
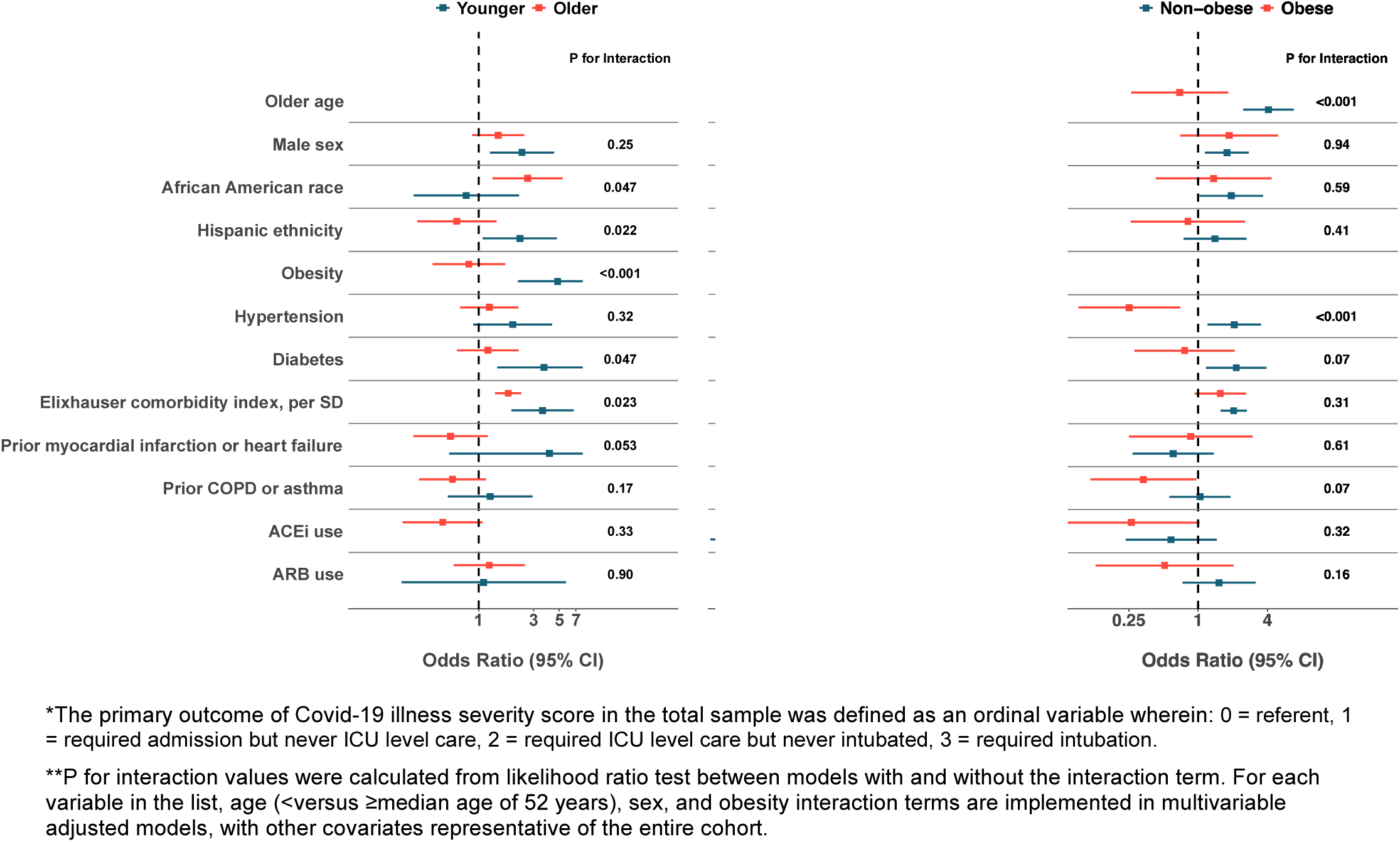
Associations with Overall Covid-19 Illness Severity, Stratified by Subgroups. Relative risks associated with illness severity score are shown for all associations observed in the total sample (N=442), stratified by subgroups defined by age (younger vs. older than median age 52 years), sex, and obesity (BMI ≥30 kg/m^2^).

## CONCLUSIONS

We examined the pre-existing characteristics associated with severity of Covid-19 illness, as observed thus far in our healthcare system located in Los Angeles, California. We found that almost half of patients presenting for evaluation and then confirmed to have Covid-19 were clinically assessed to require hospital admission. In analyses adjusting for concomitant factors, these higher risk individuals were more likely to be older, male, African American, obese, and have diabetes mellitus in addition to a greater overall burden of medical comorbidities. Notably, chronic use of an ACE inhibitor appeared related to lower illness severity, in the absence of a similar finding for ARB use. Among all individuals requiring inpatient care for Covid-19, male patients had a greater than 2.5-fold odds of needing intensive care and a 3.0-fold odds of needing intubation even after accounting for co-existing risk factors and chronic medical conditions.

Recognizing that patients with Covid-19 illness can present with clinical features that vary based on timing of the index encounter, we sought to identify the specific pre-existing traits that predispose some individuals to the more severe forms of Covid-19 illness – even after adjusting for co-existing factors. In our U.S. based metropolitan community, we observed that both obesity and diabetes mellitus are associated with a greater odds of needing hospital admission for Covid-19 but not of requiring further escalation of care; this finding is consistent with emerging reports of obesity and diabetes mellitus each being associated with a greater risk for pneumonia due to Covid-19 as well as other community-acquired viral agents – particularly in areas of the world where obesity is prevalent.^30–35^ Also consistent with worldwide reports, we observed that older age is a significant predisposing risk factor for greater Covid-19 illness severity in multivariable-adjusted models; this finding may represent an age-related immune susceptibility that is not completely captured by even a comprehensive comorbidity measure such as the Elixhauser index. Notwithstanding an overall age association in the expected direction of risk, we also found a paradoxical age interaction for certain key correlates. In effect, presence of obesity, diabetes, or an elevated overall comorbidity index were each associated with greater Covid-19 illness severity in younger (i.e. <52 years) compared to older age groups. While unexpected, this finding is actually consistent with the known reduction of ACE2 expression with advancing age, a phenomenon that has been proposed as a major contributor to the broad susceptibility to Covid-19 seen in younger to middle aged individuals across the population at large.^36^

Consistent with worldwide reports, we found that the association of male sex with greater odds for every metric of Covid-19 illness severity was especially prominent – and this was not explained by age variation, risk factors, or comorbidities.^37^ Reasons for the male predominance of illness severity remain unclear. Although ACE2 genetic expression is on the X chromosome, evidence to date would suggest relatively comparable expression levels between sexes,^38,39^ albeit with some potential for variation in relation to differences in sex hormones; select animal studies have shown increased ACE2 activity in the setting of ovariectomy and the opposite effect with orchietomy.^40,41^ While there remains scant data currently available to explain sex differences for Covid-19, male sex bias was also observed for SARS and MERS.^42,43^ Similar to the findings in our study, this increase risk was not attributable to a greater prevalence of smoking among men. Notably, prior murine studies have also demonstrated male versus female bias in susceptibility to SARS-CoV infection, which may be related to the effects of sex-specific steroids and *X*-linked gene activity on modulation of both the innate and adaptive immune response to viral infection.^44^ Further research specific to the sexual dimorphism seen in SARS-CoV-2 susceptibility is needed.

In our U.S. based metropolitan community, we also observed racial and ethnic patterns of susceptibility to greater Covid-19 illness severity. Specifically, we found that African Americans were at greater risk for needing higher levels of care overall, and this vulnerability appeared more pronounced in older age and among men. Although an overall risk association was not seen for Hispanic ethnicity, there was a trend towards greater Covid-19 illness severity in younger aged compared to older aged Hispanic/Latino persons. A recent national report from the CDC also suggests overall higher rates of Covid-19 susceptibility in African Americans, and our findings confirm this trend exists even after adjusting for age, risk factors, and comorbidities.^45^ In addition to the effects of unmeasured socioeconiomic and healthcare access variables, racial/ethnic disparities in Covid-19 illness severity may relate to yet unidentified host-viral susceptibility factors that could also be contributing to heterogeneity of community transmission seen across regions worldwide and populations at large.^46^

The use of ACE inhibitor or angiotensin receptor blocker (ARB) medications has been controversial given the possibility that these agents upregulate the expression of ACE2, which is the viral point of entry into cells.^47^ Indeed, the spike protein of the SARS-CoV-2 virus is now known to bind ACE2 to enter alveolar type 2 epithelial cells, in particular.^48^ Other data suggest that reduced ACE2 activity might lead to local renin-angiotensin-aldosterone system activation and, thus, a potential benefit for renin-angiotensin-aldosterone system inhibitors has been proposed.^36,47,49^ Although we observed a non-significant trend in association of chronic ACE inhibitor treatment with lower Covid-19 illness severity, we found evidence of neither risk nor benefit with ARBs. In future larger studies, more ideally powered to detect such associations, a protective effect of ACE inhibition on Covid-19 outcomes would raise the specter of high-level homology between ACE and ACE2, potentially allowing for ACE inhibitors to bind ACE2 and block viral entry despite the lack of inhibition of the catalytic domain.^50^ Such findings would underscore the importance of considering modified or tailored ACE inhibition to exploit the structural basis of the binding interaction between SARS-CoV-2 and ACE2.^51,52^ Together, our findings to date are supportive of current recommendations to not discontinue chronic ACE inhibitor or ARB therapy for patients with appropriate indications for these medications.

Several limitations of our study merit consideration. Our cohort included all individuals who underwent laboratory testing for Covid-19 and not individuals who did not undergo testing; thus, our study results are derived from individuals presenting with symptoms that were deemed severe enough to warrant testing. All data including past medical history data were collected from the EHR and, thus, subject to coding bias and variations in reporting quality. To minimize the potential effects of these limitations that are inherent to EHR data, we performed iterative quality checks on the dataset and conducted manual chart review to verify values for key variables. We recognize that the illness severity outcomes defined as clinically ascertained need for hospital admission, ICU level care, and intubation, may vary from practice to practice. As in many other U.S. medical centers affected by the Covid-19 pandemic, our clinical staff have been practicing under institutional guidance to conserve resources and we anticipate that the thresholds for escalating care are likely comparable; thresholds for admission, transfer to intensive care, and intubation may be different in more resource constrained environments. Given the relatively small number of observed in-hospital deaths (N=11), and thus limited statistical power to detect assocations, we deferred analyses of pre-existing characteristics and mortality risk to future investigations. The modest size of this early analysis of our growing clinical cohort may have limited the ability to detect potential additional predictors of Covid-19 illness severity; thus, further investigations are needed in larger sized samples. Finally, our results are derived from a single healthcare system, albeit multi-center and serving a large catchment of the diverse population of Los Angeles, California. Additional studies are needed to examine the extent to which our findings are generalizable to other populations affected by Covid-19.

In summary, we found that among patients tested and managed for laboratory confirmed Covid-19 in our healthcare system to date, approximately half require admission for inpatient hospital care. After accounting for the presence of concomitant risk factors, certain traits increase the odds of developing more severe forms of Covid-19 illness: older age, male sex, African American race, obesity, diabetes mellitus, and a greater overall burden of medical comorbidities. Well over a third of hospitalized patients require intensive care, with a substantial proportion needing intubation and mechanical ventilation for respiratory failure. Among hospitalized patients, the highest risk individuals were more likely to be predominantly men of any age or race – for reasons not explained by comorbidities. Further investigations are needed to understand the mechanisms underlying these associations and, in turn, determine the most optimal approaches to attenuating adverse outcomes for all persons at risk.

## Data Availability

The data that support the findings of this study are available from Cedars-Sinai Medical Center, upon reasonable request. The data are not publicly available due to the contents including information that could compromise research participant privacy/consent.

## ACKNOWLEDGEMENTS

We are grateful to all the front-line healthcare workers in our healthcare system who continue to be dedicated to delivering the highest quality care for all patients.

## SUPPLEMENTARY APPENDIX

Pre-Existing Traits Associated with Covid-19 Illness Severity

**Supplemental Table 1.**
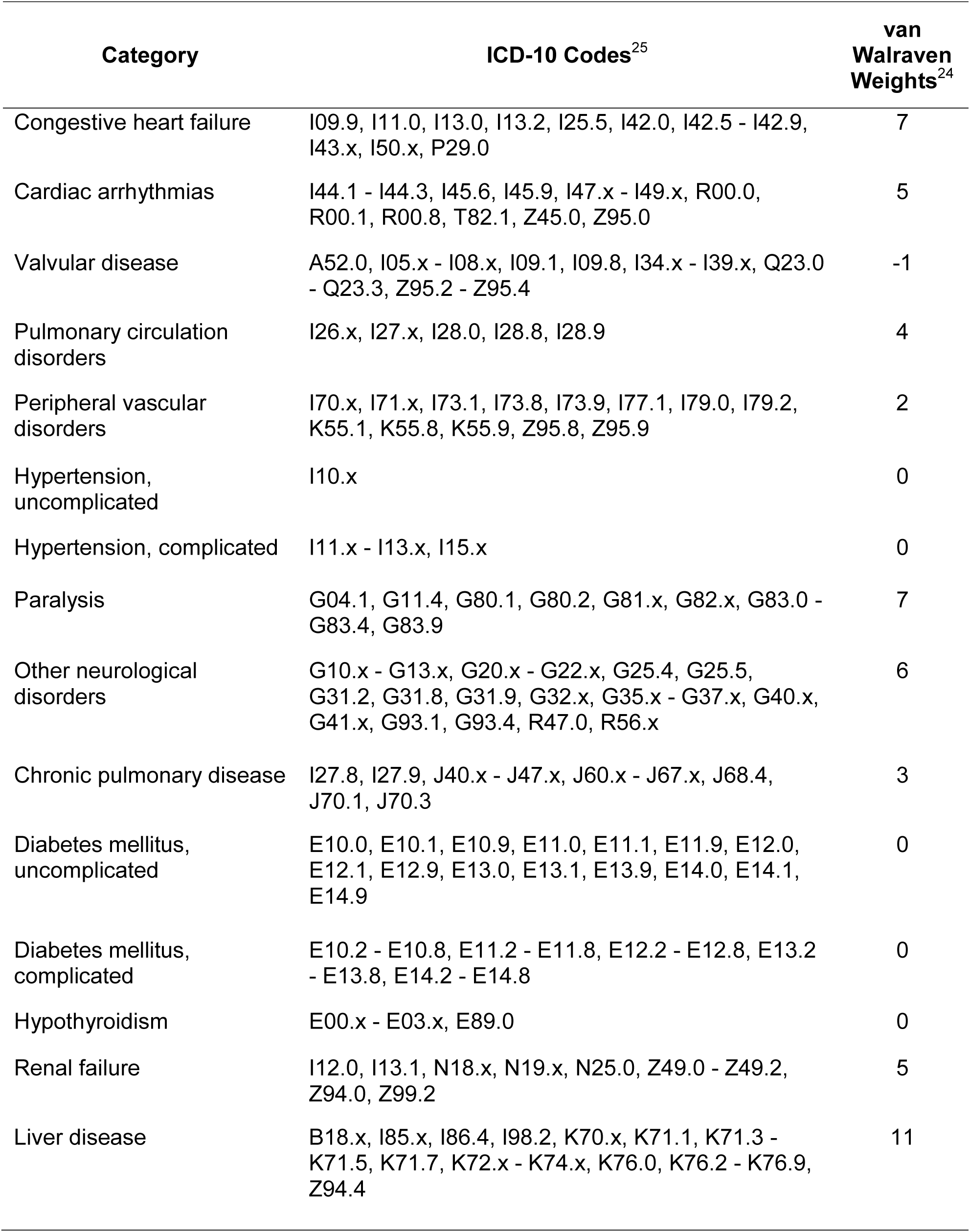
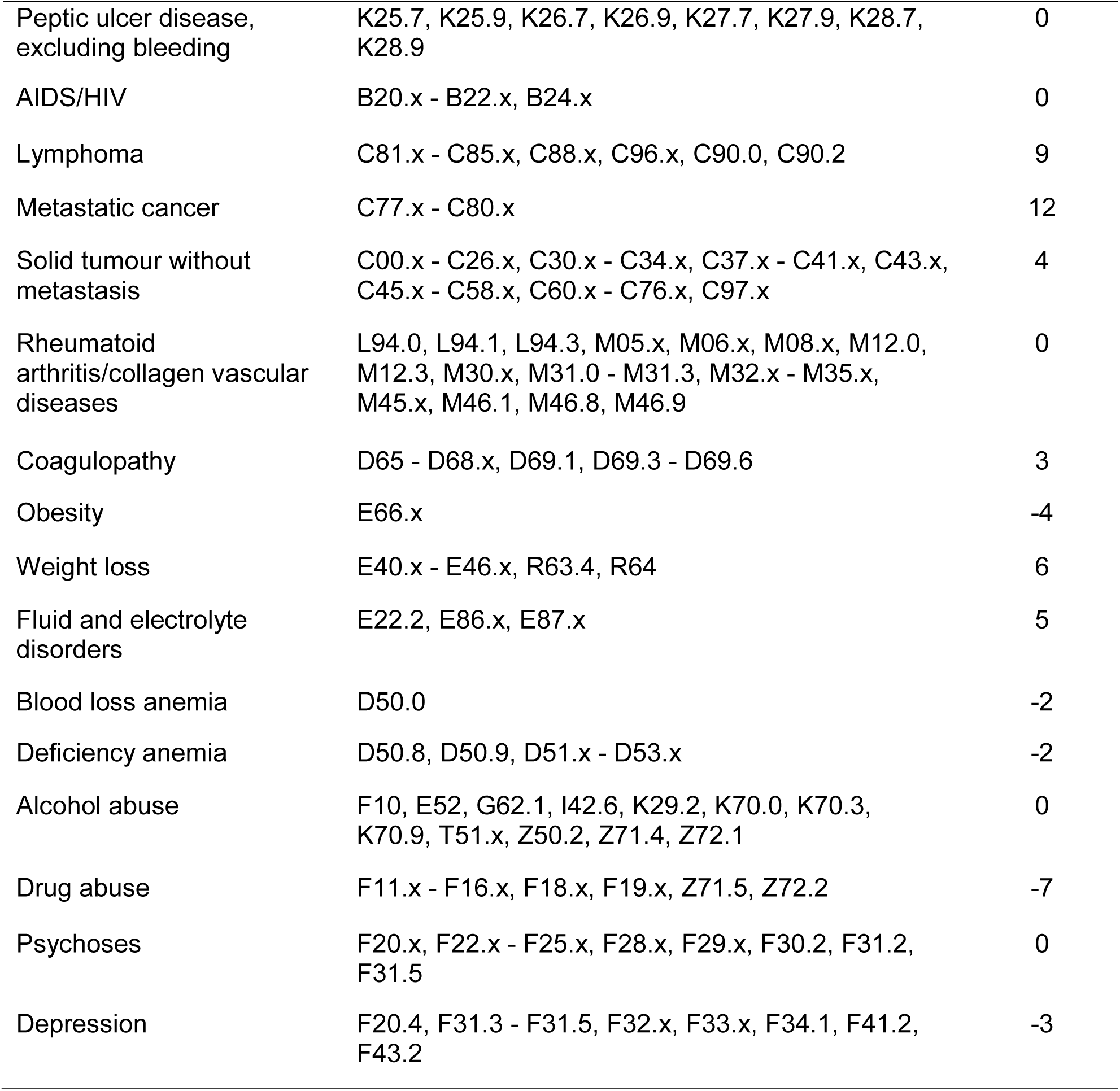
Elixhauser Comorbiditiy Index and Van Walraven Weights.

| **Category** | **ICD-10 Codes<sup>25</sup>** | **van Walraven Weights<sup>24</sup>** |
| --- | --- | --- |
| Congestive heart failure | I09.9, I11.0, I13.0, I13.2, I25.5, I42.0, I42.5 - I42.9, I43.x, I50.x, P29.0 | 7 |
| Cardiac arrhythmias | I44.1 - I44.3, I45.6, I45.9, I47.x - I49.x, R00.0, R00.1, R00.8, T82.1, Z45.0, Z95.0 | 5 |
| Valvular disease | A52.0, I05.x - I08.x, I09.1, I09.8, I34.x - I39.x, Q23.0 - Q23.3, Z95.2 - Z95.4 | -1 |
| Pulmonary circulation disorders | I26.x, I27.x, I28.0, I28.8, I28.9 | 4 |
| Peripheral vascular disorders | I70.x, I71.x, I73.1, I73.8, I73.9, I77.1, I79.0, I79.2, K55.1, K55.8, K55.9, Z95.8, Z95.9 | 2 |
| Hypertension, uncomplicated | I10.x | 0 |
| Hypertension, complicated | I11.x - I13.x, I15.x | 0 |
| Paralysis | G04.1, G11.4, G80.1, G80.2, G81.x, G82.x, G83.0 - G83.4, G83.9 | 7 |
| Other neurological disorders | G10.x - G13.x, G20.x - G22.x, G25.4, G25.5, G31.2, G31.8, G31.9, G32.x, G35.x - G37.x, G40.x, G41.x, G93.1, G93.4, R47.0, R56.x | 6 |
| Chronic pulmonary disease | I27.8, I27.9, J40.x - J47.x, J60.x - J67.x, J68.4, J70.1, J70.3 | 3 |
| Diabetes mellitus, uncomplicated | E10.0, E10.1, E10.9, E11.0, E11.1, E11.9, E12.0, E12.1, E12.9, E13.0, E13.1, E13.9, E14.0, E14.1, E14.9 | 0 |
| Diabetes mellitus, complicated | E10.2 - E10.8, E11.2 - E11.8, E12.2 - E12.8, E13.2 - E13.8, E14.2 - E14.8 | 0 |
| Hypothyroidism | E00.x - E03.x, E89.0 | 0 |
| Renal failure | I12.0, I13.1, N18.x, N19.x, N25.0, Z49.0 - Z49.2, Z94.0, Z99.2 | 5 |
| Liver disease | B18.x, I85.x, I86.4, I98.2, K70.x, K71.1, K71.3 - K71.5, K71.7, K72.x - K74.x, K76.0, K76.2 - K76.9, Z94.4 | 11 |
| Peptic ulcer disease, excluding bleeding | K25.7, K25.9, K26.7, K26.9, K27.7, K27.9, K28.7, K28.9 | 0 |
| AIDS/HIV | B20.x - B22.x, B24.x | 0 |
| Lymphoma | C81.x - C85.x, C88.x, C96.x, C90.0, C90.2 | 9 |
| Metastatic cancer | C77.x - C80.x | 12 |
| Solid tumour without metastasis | C00.x - C26.x, C30.x - C34.x, C37.x - C41.x, C43.x, C45.x - C58.x, C60.x - C76.x, C97.x | 4 |
| Rheumatoid arthritis/collagen vascular diseases | L94.0, L94.1, L94.3, M05.x, M06.x, M08.x, M12.0, M12.3, M30.x, M31.0 - M31.3, M32.x - M35.x, M45.x, M46.1, M46.8, M46.9 | 0 |
| Coagulopathy | D65 - D68.x, D69.1, D69.3 - D69.6 | 3 |
| Obesity | E66.x | -4 |
| Weight loss | E40.x - E46.x, R63.4, R64 | 6 |
| Fluid and electrolyte disorders | E22.2, E86.x, E87.x | 5 |
| Blood loss anemia | D50.0 | -2 |
| Deficiency anemia | D50.8, D50.9, D51.x - D53.x | -2 |
| Alcohol abuse | F10, E52, G62.1, I42.6, K29.2, K70.0, K70.3, K70.9, T51.x, Z50.2, Z71.4, Z72.1 | 0 |
| Drug abuse | F11.x - F16.x, F18.x, F19.x, Z71.5, Z72.2 | -7 |
| Psychoses | F20.x, F22.x - F25.x, F28.x, F29.x, F30.2, F31.2, F31.5 | 0 |
| Depression | F20.4, F31.3 - F31.5, F32.x, F33.x, F34.1, F41.2, F43.2 | -3 |

**Supplemental Table 2.** Characteristics Associated with Need for Any Hospitalization in All Patients with Covid-19.

|  | Age- and Sex-Adjusted Models |  | Multivariable-Adjusted Model* |  |
| --- | --- | --- | --- | --- |
|  | OR (95% CI) | *P* | OR (95% CI) | *P* |
| **Outcome: Severe Illness (N=214 needed any hospital admission, of the N=442 total diagnosed with COVID-19)** |  |  |  |  |
| **Age, per 10 years** | 1.88 (1.65,2.14) | <0.001 | **1.55 (1.32,1.81)** | **<0.001** |
| Male sex | 1.62 (1.04,2.52) | 0.034 | 1.64 (0.99,2.72) | 0.054 |
| African American race | 2.30 (1.18,4.50) | 0.015 | 1.66 (0.78,3.53) | 0.19 |
| Hispanic ethnicity | 1.65 (0.91,2.99) | 0.10 | 1.16 (0.58,2.33) | 0.67 |
| Obesity | 2.04 (1.14,3.65) | 0.016 | 1.99 (0.97,4.08) | 0.059 |
| Hypertension | 2.09 (1.27,3.44) | 0.004 | 1.14 (0.61,2.13) | 0.69 |
| **Diabetes mellitus** | 3.37 (1.82,6.24) | <0.001 | **2.81 (1.35,5.85)** | **0.006** |
| **Elixhauser comorbidity score, per SD** | 4.27 (2.64,6.90) | <0.001 | **4.34 (2.53,7.44)** | **<0.001** |
| Prior myocardial infarction or heart failure | 5.11 (1.71,15.28) | 0.004 | 0.69 (0.17,2.82) | 0.60 |
| Prior COPD or asthma | 1.53 (0.83,2.80) | 0.17 | 0.80 (0.38,1.65) | 0.54 |
| ACE inhibitor use | 0.79 (0.34,1.84) | 0.58 | 0.42 (0.14,1.25) | 0.12 |
| Angiotensin receptor blocker use | 0.90 (0.41,1.95) | 0.78 | 0.85 (0.34,2.09) | 0.72 |

\*All listed covariates shown were included in the full multivariable-adjusted model.

**Supplemental Table 3.** Characteristics Associated with Distinct Outcomes in Patients Hospitalized for Covid-19.

|  | Age- and Sex-Adjusted Models |  | Multivariable-Adjusted Model* |  |
| --- | --- | --- | --- | --- |
|  | OR (95% CI) | P | OR (95% CI) | P |
| **Outcome: Critical Illness (N=77 needing ICU level care, of N=214 admitted)** |  |  |  |  |
| Age, per 10 years | 1.14 (0.97,1.33) | 0.11 | 1.11 (0.93,1.32) | 0.25 |
| Male sex | 2.29 (1.23,4.26) | 0.009 | **2.41 (1.27,4.55)** | **0.007** |
| African American race | 1.75 (0.85,3.60) | 0.13 | 1.51 (0.72,3.18) | 0.28 |
| Hispanic ethnicity | 1.27 (0.58,2.78) | 0.55 |  |  |
| Obesity | 1.26 (0.62,2.57) | 0.52 | - | - |
| Hypertension | 1.56 (0.83,2.92) | 0.17 | 1.62 (0.84,3.11) | 0.15 |
| Diabetes mellitus | 1.18 (0.64,2.19) | 0.59 | - | - |
| Elixhauser comorbidity score, per SD | 1.12 (0.86,1.47) | 0.39 | - | - |
| Prior myocardial infarction or heart failure | 1.07 (0.52,2.23) | 0.85 | - | - |
| Prior COPD or asthma | 0.82 (0.40,1.68) | 0.59 | - | - |
| ACE inhibitor use | 0.43 (0.14,1.26) | 0.12 | 0.38 (0.13,1.17) | 0.09 |
| Angiotensin receptor blocker use | 1.47 (0.64,3.38) | 0.37 | - | - |
| **Outcome: Respiratory Failure (N=52 needing intubation, of N=214 admitted)** |  |  |  |  |
| Age, per 10 years | 1.15 (0.96,1.37) | 0.14 | 1.14 (0.95,1.38) | 0.18 |
| Male sex | 2.88 (1.36,6.06) | 0.005 | 2.99 (1.41,6.36) | 0.004 |
| African American race | 2.14 (0.99,4.64) | 0.053 | 2.14 (0.99,4.64) | 0.053 |
| Hispanic ethnicity | 1.35 (0.56,3.24) | 0.51 | - | - |
| Obesity | 1.57 (0.72,3.41) | 0.26 | - | - |
| Hypertension | 0.81 (0.40,1.64) | 0.57 | - | - |
| Diabetes mellitus | 0.94 (0.47,1.89) | 0.87 | - | - |
| Elixhauser comorbidity score, per SD | 0.86 (0.63,1.18) | 0.34 | - | - |
| Prior myocardial infarction or heart failure | 0.70 (0.30,1.64) | 0.41 | - | - |
| Prior COPD or asthma | 0.67 (0.29,1.53) | 0.34 | - | - |
| ACE inhibitor use | 0.57 (0.17,1.85) | 0.35 | - | - |
| Angiotensin receptor blocker use | 1.38 (0.56,3.42) | 0.48 | - | - |

\*To avoid model overfitting given the sample size, covariates included in the multivariable model were selected from age- and sex-adjusted models based on significance with $P < 0.20$ .

**Supplemental Table 4.** Age, Sex, and Body Mass Index Interactions With Characteristics Associated with Overall Covid-19 Illness Severity in the Total Sample.

|  | Interaction with Age Group |  | Interaction with Male Sex |  | Interaction with Obesity |  |
| --- | --- | --- | --- | --- | --- | --- |
|  | LRT Chi-square | *P value* | LRT Chi-square | *P value* | LRT Chi-square | *P value* |
| Older age (≥52 years) | - | - | 1.31 | 0.25 | 10.97 | **<0.001** |
| Male sex | 1.31 | 0.25 | - | - | 0.01 | 0.94 |
| African American race | 3.95 | **0.047** | 3.38 | 0.07 | 0.30 | 0.59 |
| Hispanic ethnicity | 5.27 | **0.022** | 0.01 | 0.93 | 0.68 | 0.41 |
| Obesity | 10.97 | **<0.001** | 0.01 | 0.94 | - | - |
| Hypertension | 0.99 | 0.32 | 0.42 | 0.52 | 14.73 | **<0.001** |
| Diabetes mellitus | 3.96 | **0.047** | 0.38 | 0.54 | 3.23 | 0.07 |
| Elixhauser comorbidity score, per SD | 5.15 | **0.023** | 0.03 | 0.86 | 1.01 | 0.31 |
| Prior myocardial infarction or heart failure | 3.73 | 0.053 | 0.00 | 0.97 | 0.26 | 0.61 |
| Prior COPD or asthma | 1.86 | 0.17 | 0.35 | 0.56 | 3.40 | 0.07 |
| ACE inhibitor use | 0.97 | 0.33 | 0.50 | 0.48 | 0.97 | 0.32 |
| Angiotensin receptor blocker use | 0.02 | 0.90 | 0.22 | 0.64 | 1.97 | 0.16 |

\*The primary outcome of Covid-19 illness severity score in the total sample was defined as an ordinal variable wherein: 0 = referent, 1 = required admission but never ICU level care, 2 = required ICU level care but never intubated, 3 = required intubation.

**Supplemental Table 5.**
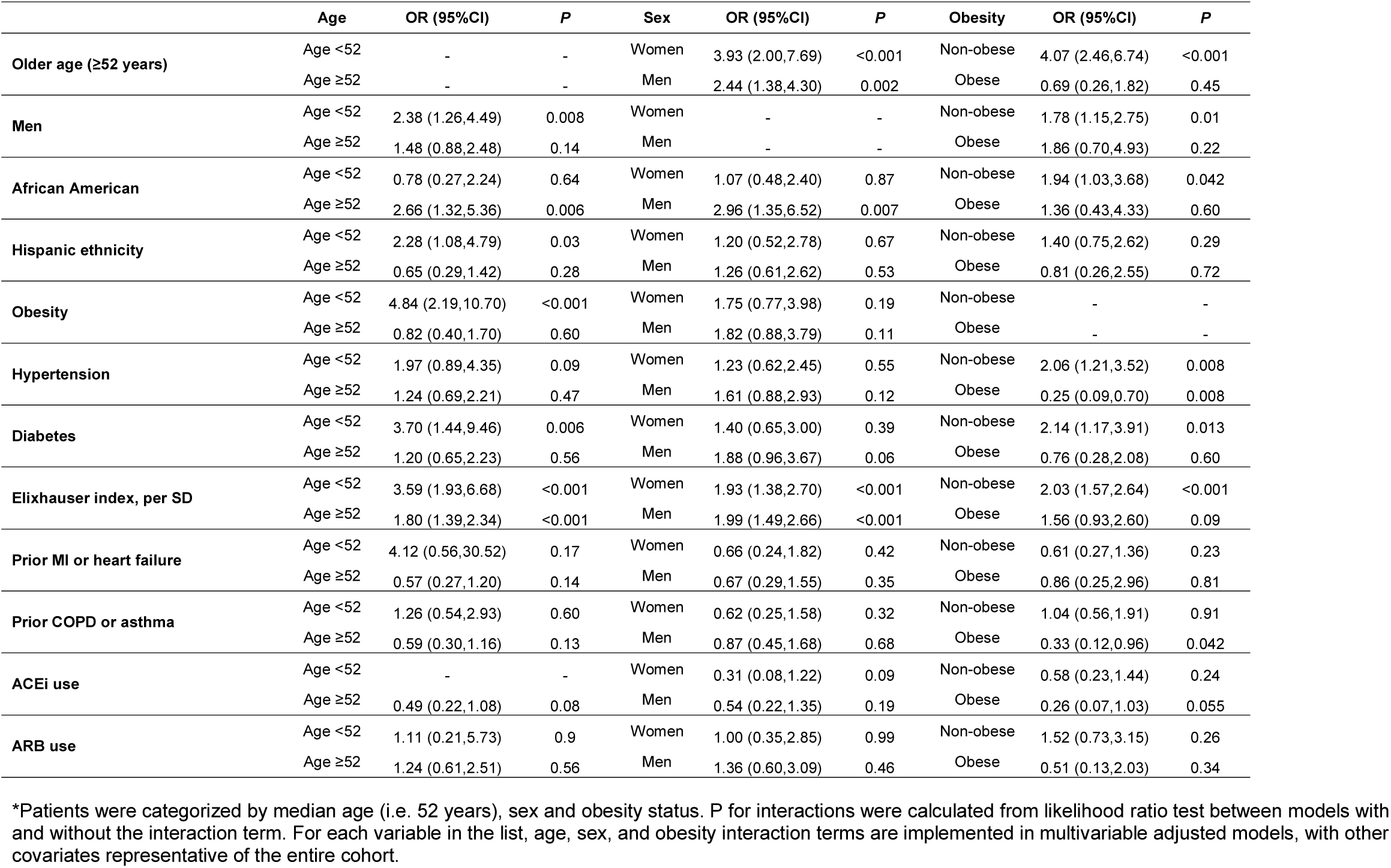
Age, Sex, and Obesity Stratified Associations with Overall Covid-19 Illness Severity in the Total Sample.

**Supplemental Figure 1.**
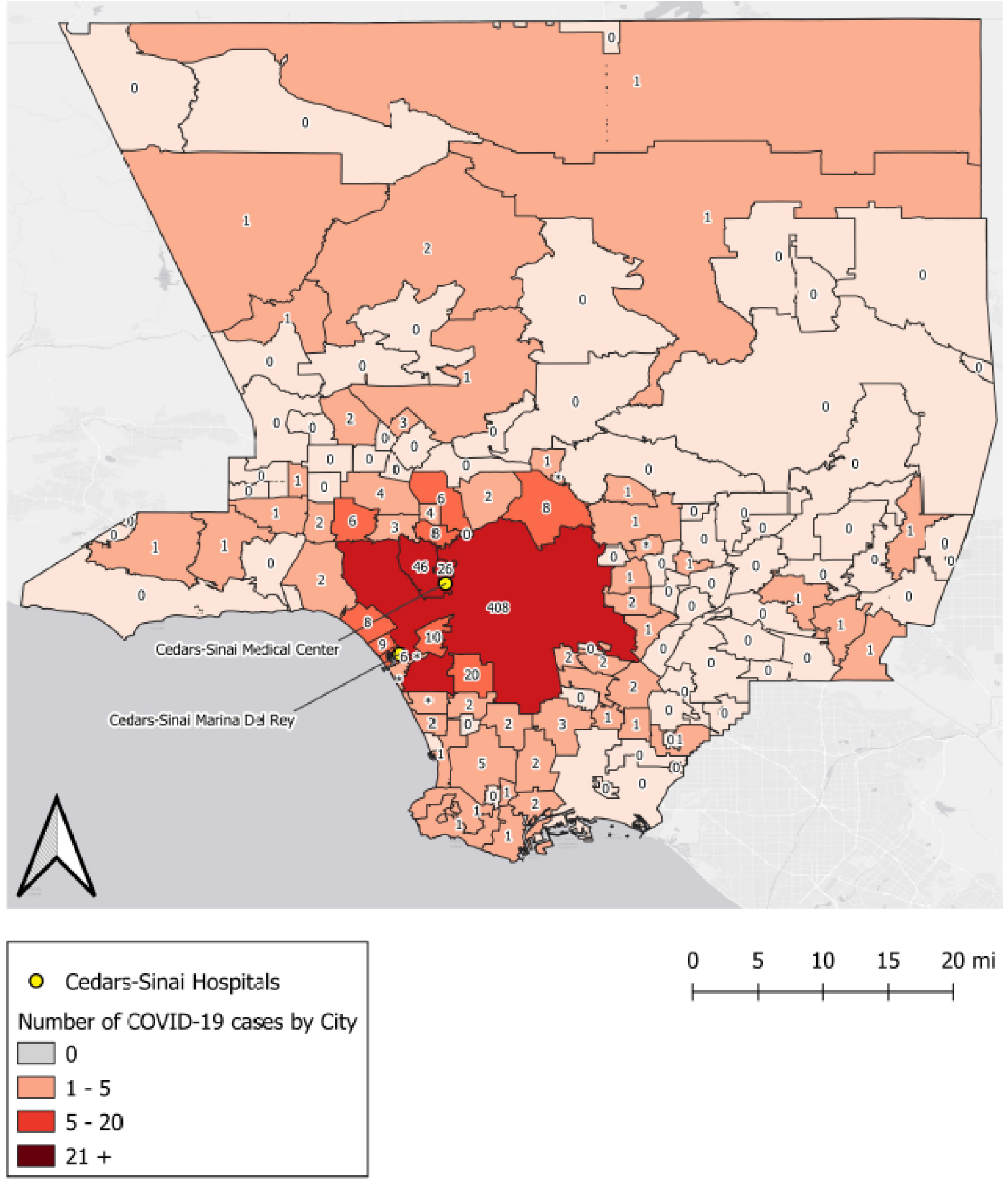
Los Angeles County Regional Distribution of All Patients with Covid-19. The patients treated in our healthcare system for Covid-19 illness presented from across a diverse regional distribution of residential locations across Los Angeles County.

